# Quantitative susceptibility mapping at 7 Tesla in COVID-19: mechanistic and outcome associations

**DOI:** 10.1101/2023.11.22.23298899

**Authors:** Catarina Rua, Betty Raman, Christopher T Rodgers, Virginia FJ Newcombe, Anne Manktelow, Doris A Chatfield, Stephen J. Sawcer, Joanne G Outtrim, Victoria C Lupson, Emmanuel A Stamatakis, Guy B Williams, William T Clarke, Lin Qiu, Martyn Ezra, Rory McDonald, Stuart Clare, Mark Cassar, Stefan Neubauer, Karen D Ersche, Edward T Bullmore, David K Menon, Kyle Pattinson, James B. Rowe, the Cambridge NeuroCOVID group and the CITIID-NIHR COVID-19 BioResourceCollaboration

## Abstract

*Post mortem* studies have shown that patients dying from severe SARS-CoV-2 infection frequently have pathological changes in their central nervous system, particularly in the brainstem. Many of these changes are proposed to result from para-infectious and/or post-infection immune responses. Clinical symptoms such as fatigue, breathlessness, and chest pain are frequently reported in post-hospitalized COVID-19 patients. We propose that these symptoms are in part due to damage to key neuromodulatory brainstem nuclei. While brainstem involvement has been demonstrated in the acute phase of the illness, the evidence of long-term brainstem change on magnetic resonance imaging (MRI) is inconclusive. We therefore used ultra-high field (7T) quantitative susceptibility mapping (QSM) to test the hypothesis that brainstem abnormalities persist in post-COVID patients and that these are associated with persistence of key symptoms.

We used 7T QSM data from 30 patients, scanned 93 – 548 days after hospital admission for COVID-19 and compared them to 51 age-matched controls without prior history of COVID-19 infection. We correlated the patients’ QSM signals with disease severity (duration of hospital admission and COVID-19 severity scale), inflammatory response during the acute illness (C-reactive protein, D-Dimer and platelet levels), functional recovery (modified Rankin scale; mRS), depression (PHQ-9) and anxiety (GAD-7).

In COVID-19 survivors the MR susceptibility increased in the medulla, pons and midbrain regions of the brainstem. Specifically, there was increased susceptibility in the inferior medullary reticular formation and the raphe pallidus and obscurus. In these regions, patients with higher tissue susceptibility had worse acute disease severity, higher acute inflammatory markers, and significantly worse functional recovery.

Using non-invasive ultra-high field 7T MRI, we show evidence of brainstem pathophysiological changes associated with inflammatory processes in post-hospitalized COVID-19 survivors. This study contributes to understanding the mechanisms of long-term effects of COVID-19 and recovery.

## 1. Introduction

Neuroradiological changes have been described in severely affected hospitalised patients with severe acute respiratory syndrome coronavirus (SARS-CoV-2) causing coronavirus disease of 2019 (COVID-19). The most common acute findings are cerebral microhemorrhages, encephalopathy and white matter hyperintensities.^1–9^ Brainstem involvement in COVID-19 has also been reported in autopsy studies^10,11^, which show tissue neurodegeneration and inflammatory responses. These abnormalities are reflected by MRI-visible changes in the brainstem in the acute phase of the illness.^8^ Indeed, such brainstem abnormalities have been proposed^12,13^ as a mechanism for the Post-Acute Covid Syndrome 2 (PACS) which may be related to “long-COVID”.^14^ This syndrome includes somatic symptoms (such as fatigue and breathlessness, often in the absence of objectively demonstrable cardiorespiratory abnormalities), cognitive deficits (sometimes referred to as “brain fog”), and mental health problems (such as anxiety, depression, and post-traumatic stress disorder). However, conventional 3T MRI has not shown consistent brainstem abnormalities at follow up. More advanced MRI techniques such as quantitative susceptibility mapping (QSM) have potential to identify more subtle abnormalities, which could reveal neuroanatomical changes in the brainstem after COVID infection.

*In vivo* QSM is a post-processing technique applied to T_2_*-weighted gradient-echo (GRE) images. Constituents of tissue can contribute a negative (or “diamagnetic”) susceptibility (e.g. soft tissue, calcium, myelin) or a positive (or “paramagnetic”) susceptibility (e.g. iron, aluminium, copper). QSM is effective to detect cerebral microbleeds^15^, increases in iron deposition in the basal ganglia and midbrain with age and in disease^16–18^, to differentiate calcified from haemorrhagic lesions^19^, and to detect chronic inflammation in multiple sclerosis.^20^ Furthermore, high-resolution, ultra-high field (≥7T) QSM has improved susceptibility contrast in cortical and subcortical tissues^21^, which provide greater sensitivity to detect microstructural alterations.

By capitalising on a preliminary analysis showing abnormal brainstem QSM in post-hospitalised patients with COVID-19^22^, we proceeded to investigate QSM abnormalities in the brainstem, according to sub-regions (midbrain, pons, medulla and superior cerebellar peduncle (SCP)) defined *a priori* as regions-of-interest (ROI). Furthermore, to increase regional specificity, we also performed the group analysis on a voxel-by-voxel approach to localize specific clusters in the brainstem showing atrophy. Finally, we tested whether susceptibility in the brainstem correlates with clinical measures of disease severity, laboratory measures of inflammation, and measures of recovery with similar regional-wise and voxel-wise analysis.

## 2. Methods

### 2.1 Participants

We recruited people who were hospitalised with COVID-19 and subsequently discharged as “post-hospitalized patients” (COVID group; N=31, 18 males, age 57±12 y.o.) for scanning by 7T MRI at two sites: (1) site-1 at the Wolfson Brain Imaging Centre (WBIC, Cambridge, UK) and (2) site-2 at the Wellcome Centre for Integrative Neuroimaging (WIN, Oxford, UK) (Supplementary Information 1). Inclusion criteria were: (1) evidence of COVID-19 infection confirmed by SARS-CoV-2 polymerase chain reaction (PCR) of respiratory samples (nasal or throat swab), (2) no specific pre-COVID history of neurological or psychiatric disorders, and (3) no contradictions to 7T MRI.

COVID-19 severity was determined during hospital admission using the WHO ordinal scale for clinical improvement.^23^ Peak CRP and D-Dimer levels, and lowest platelet levels during hospital stay were recorded. At the time of the follow up clinic (time between clinic and imaging was 48±21 days for site-1 and 115±34 for site-2), functional recovery was assessed using the modified Rankin scale (mRS), and mental health was assessed using two sets of questionnaires for anxiety and depression, respectively, the Generalised Anxiety Disorder-7 (GAD-7) and the Patient Health Questionnaire-9 (PHQ-9).

Healthy controls (HC) were scanned by 7T MRI in site-1 (HC group, N=51, 34 males, age 53±15 y.o.). These came from two subgroups: people scanned before December 2019 i.e. before possible exposure to COVID-19 (“HC1 subgroup”); and people scanned during the pandemic before April 2021 who were asymptomatic with no history of positive SARS-CoV-2 PCR (“HC2 subgroup”) (Supplementary Information 1).

The study was approved by the following ethics committees: Cambridgeshire Research Ethics Committee HBREC.2016.13.am3, East of England Research Ethics Committee 17/EE/0025 , Norfolk Research Ethics Committee EE/0395, and North West Preston

Research Ethics Committee 20/NW/0235. All participants provided informed consent in accordance with the Declaration of Helsinki.

### 2.2 MRI acquisition

All participants had 7T MRI using a 32-channel head coil (Nova Medical, USA). Site-1 used their 7T Terra scanner (Siemens, Germany) and Site-2 used their Magnetom 7T (Siemens, Germany). Following previous published results on the reproducibility across these two scanners for QSM^24^, both sites acquired 3D T_2_*-weighted (T_2_*w) multi-echo gradient-echo with 0.7mm isotropic voxels, 4.68ms TE1, 27ms TR, 6 echoes, 3.24msecho spacing, 15° nominal flip angle, 430Hz/pixel bandwidth, 2×2 acceleration-factor, over a 224×196×157mm^3^ field of view. MP2RAGE T_1_-weighted (T_1_w) scans were acquired for anatomical localization and registration. For the COVID and HC2 groups this used the UK7T harmonized protocol^25^: 0.7mm isotropic voxels, 2.64ms echo time, 3500ms TR, 300 Hz/pixel bandwidth, 725/3150ms TI, 5°/2° nominal flip angles and 224×224×224 matrix. For the HC1 group we used: 0.75mm isotropic voxels, 1.99ms TE, 4300ms TR, 250Hz/pixel bandwidth, 840/2370ms TI, 5°/6° nominal flip angles and 240×224×168 matrix.

### 2.3 Data processing

Image processing used routines from the advanced normalization tools (ANTs) v2.2.0, FMRIB software library (FSL) v6.0.1, statistical parametric mapping library (SPM12) v7219 and MATLAB R2018b. Per channel data were combined as previously described at 7T.^26^ Quantitative susceptibility (χ) maps were estimated from the coil-combined T_2_*w phase data using the multi-scale dipole inversion algorithm in QSMbox^27^, as previously described.^24^ T_1_w structural images were computed from the raw images as previously described^24^ for the HC2 and COVID groups, and using the vendor supplied method for the HC1 group. All T_1_w scans were then bias-field corrected with ANTs, segmented with SPM12 and skull-stripped. We transformed the per-subject susceptibility maps into the 0.5mm isotropic ICBM 2009b standardized space for statistical analysis as described in Supplementary Information 2.

### 2.4 Region-of-interest (ROIs) analysis

Brainstem ROIs were defined using the “-brainstem-structures” tool^28^ in FreeSurfer (v6.0.0) on the ICBM T1w image to extract ROIs for brainstem and four sub-regions: the midbrain, pons, medulla and superior cerebellar peduncle (SCP). Mean susceptibility was extracted per ROI and used for further analysis.

QSM data from both sites used matched protocols developed during the UK7T harmonization project.^24,25^ Nevertheless, we tested for possible between-site effects by fitting a linear model to susceptibility at each ROI adding site as a fixed effect, age and sex as covariates, and subject as a random effect. No significant site effects were detected (Supplementary Information 3), and hence further analyses further analyses treated the COVID data as a single group.

We fitted linear models separately for each ROI, with group (COVID versus HC) as a fixed effect allowing the intercept to vary across participants (random effect). Considering that there may be age-related trends in QSM χ^16,18^, with associated gender by age effects^29^ we added age, gender and their interaction as covariates. We report frequentist hypothesis testing results, with false discovery rate (FDR) corrected p-value<0.05 for significance, Cohen’s d, 95% confidence interval (CI). We also report Bayesian model comparisons in terms of Bayes Factor (BF) and posterior probability, with BF>3 defined per conventional criteria as evidence in favour of the alternative hypothesis and BF>20 as very strong evidence.^30^ Conversely, BF<1/3 and BF<1/20 re interested as evidence and very strong evidence for the null hypothesis respectively, which cannot be inferred from “non-significant” frequentist tests’ p-values.

### 2.5 Voxelwise analyses and association with clinical and laboratory outcomes

Because the brainstem and subregions showed strong group differences (see Results sub-section 3.1), an additional voxelwise analysis comparing the COVID-vs-HC groups within the brainstem ROI was undertaken to improve the resolution for spatial distributions of small cluster differences (note the emphasis was localisation of clusters in the voxel-wise analysis, not the significance of differences given the non-independence of ROI in the voxel-wise tests). The co-registered susceptibility maps were masked by the brainstem ROI and subjected to general linear models for testing group differences.

The voxelwise analysis was performed with the randomise function in FSL, setting the number of permutations to 7,000, and the threshold free cluster enhancement (TFCE) method for cluster inference. Within these analyses, models included age, gender, age by gender interaction as covariates. To isolate the most significant clusters, a conservative family-wise error (FWE) corrected p-value < 0.01 was used to determine significant voxels, and the function *cluster* in FSL was applied to group significant clusters. We report also FWE corrected p-value < 0.05 results in supplementary information, for reference. The centroid and spatial extent of the clusters were evaluated in MNI space. The brainstem Navigator Atlas (https://brainstemimaginglab.martinos.org/research/) ROIs that overlapped the significant clusters were reported for spatial identification of the cluster location.

From the patient data, we extracted the mean χ from the significant clusters (FWE threshold p-value < 0.01) and fitted nine linear models to test their association with clinical and laboratory outcomes (WHO score, period of hospital admission, highest CRP during admission, highest D-Dimer during admission, lowest platelets during admission, GAD-7, PHQ-9 and mRS). As some clinical and laboratory measurements were not available for all subjects, we performed the linear mixed effects model by dropping patient data that did not include the measurement of interest (Supplementary Information 4).

Each model also included age, gender, age by gender interaction, time from hospital admission to scan, and cluster number as fixed effects, and subject as random effect. The Shapiro-wilk test was used to test normality of the outcome variable, and the model fit was evaluated for multicollinearity, normality distribution of residuals and homoscedasticity with the package *sjPlot* from R. When testing for the effect of period hospital admission, one subject was an outlier (hospital admission 134 days) and was excluded (Supplementary Information 5). We report both frequentist hypothesis testing results and Bayesian statistics.

## 3. Results

### 3.1 Demographic and clinical characteristics

Overall, there were no significant differences between the two groups in age, but there were more males than females in the HC group compared to the COVID group (χ^2^ = 4.92, p = 0.03). One subject from site-1 presented very low QSM signal (due to lack of signal in the brainstem, Supplementary Information 6) and was excluded in analysis. Consequently, data from fifty-one healthy controls and thirty patients were used for further analysis. The demographic and clinical features of participants used in the analyses are shown in Table 1. On the COVID group, the median time from hospital admission to the MRI scan was 199 days.

**Table 1.**
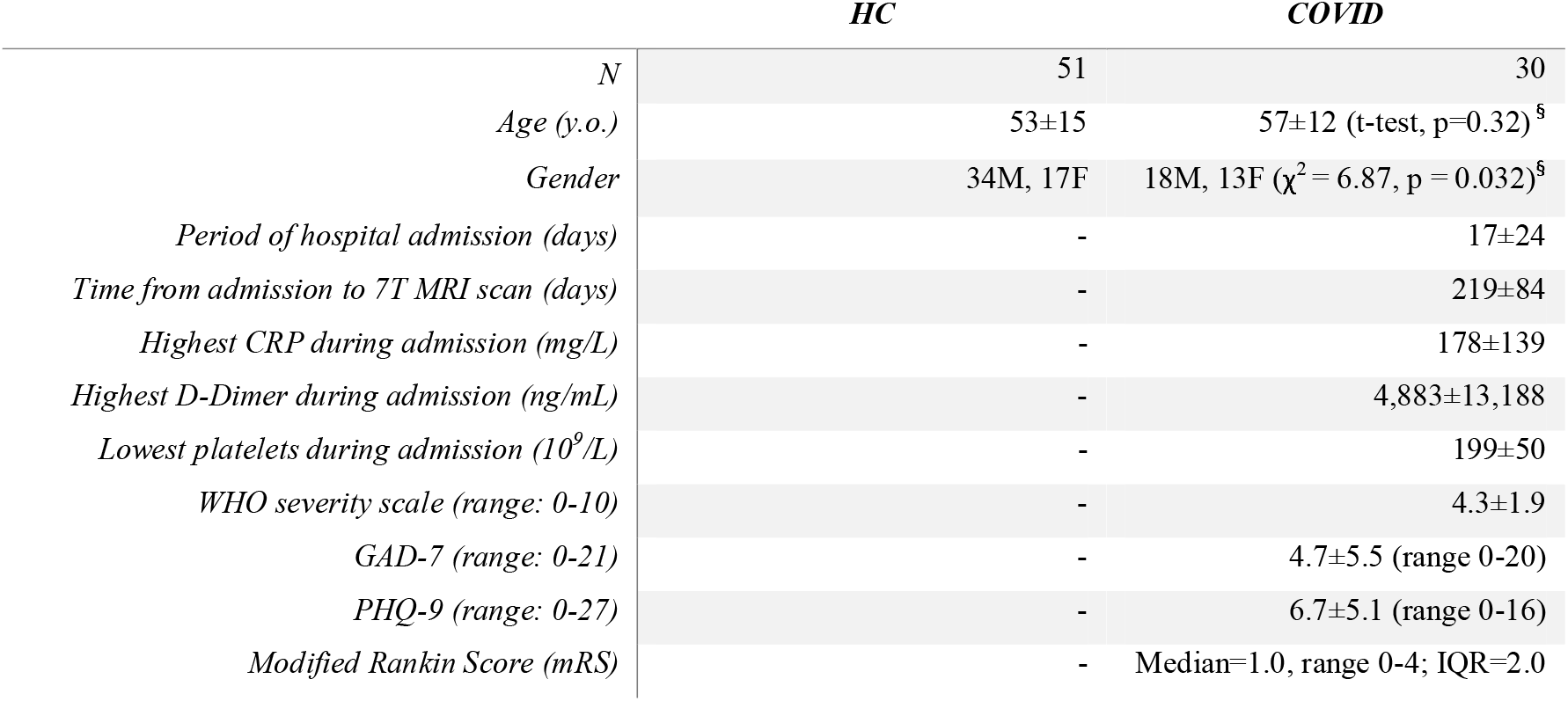
Demographics, clinical and scan data of the subjects used in the analysis of this study. Within-group mean ± standard deviation values reported when appropriate. Group-1 and group-2 are healthy control datasets from two clinical projects selected due to having identical imaging sequences and comparable spread of ages. Disease severity, blood serum values and clinical assessments are reported for the COVID-19 group. ^§^Tests comparing the HC and COVID groups.

|  | HC | COVID |
| --- | --- | --- |
| *N* | 51 | 30 |
| *Age (y.o.)* | 53±15 | 57±12 (t-test, p=0.32) <sup>§</sup> |
| *Gender* | 34M, 17F | 18M, 13F ($\chi^2 = 6.87$, p = 0.032) <sup>§</sup> |
| *Period of hospital admission (days)* | - | 17±24 |
| *Time from admission to 7T MRI scan (days)* | - | 219±84 |
| *Highest CRP during admission (mg/L)* | - | 178±139 |
| *Highest D-Dimer during admission (ng/mL)* | - | 4,883±13,188 |
| *Lowest platelets during admission ($10^9/L$)* | - | 199±50 |
| *WHO severity scale (range: 0-10)* | - | 4.3±1.9 |
| *GAD-7 (range: 0-21)* | - | 4.7±5.5 (range 0-20) |
| *PHQ-9 (range: 0-27)* | - | 6.7±5.1 (range 0-16) |
| *Modified Rankin Score (mRS)* | - | Median=1.0, range 0-4; IQR=2.0 |

### 3.1 Group differences

The regional mean χ increased in the brainstem in the COVID group compared to the healthy control group (Figure 1, 2; p(FDR) < 0.0001, d = 4.96, 95% CI [0.0030, 0.0071], Pr(post.) = 1.00, BF = 4688). This significant increase in χ was mainly localized in the Pons sub-region of the brainstem (Figure 2, p(FDR) = 0.00042, d = 4.01, 95% CI [0.0024 0.0071], Pr(post.) = 0.99, BF = 108) and in the Medulla (Figure 2, p(FDR) = 0.0035, d = 3.37, 95% CI [0.0027, 0.010], Pr(post.) = 0.96, BF = 23). The mid brain sub-region only showed a weak significant group effect (Figure 2, p(FDR) = 0.032, d = 1.99, 95% CI [0.0000042, 0.0091], Pr(post.) = 0.69, BF = 2.27). There was no group difference in the SCP (Figure 2).

**Figure 1.**
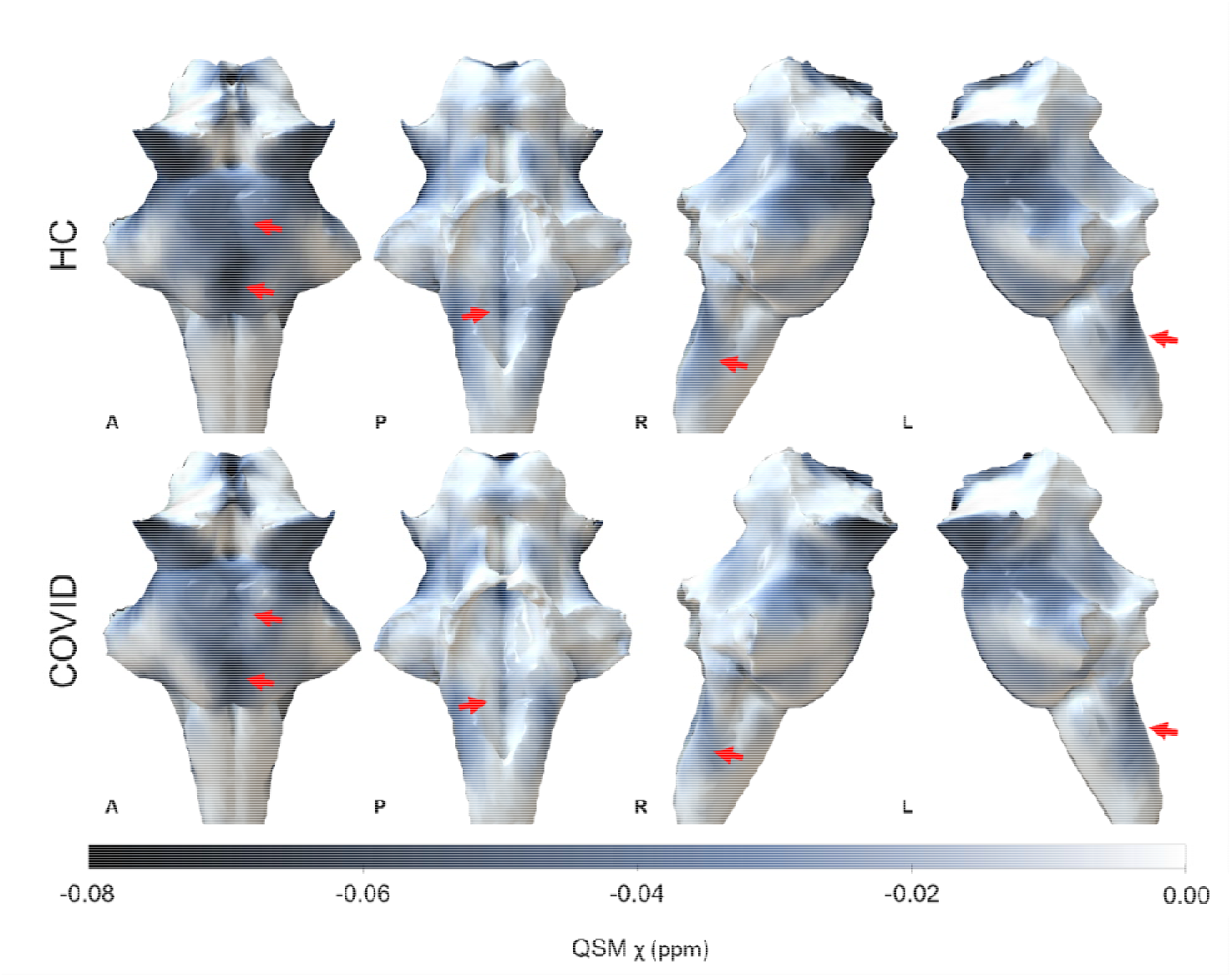
3D projections of the QSM χ maps on the rendered brainstem ROI extracted from the FreeSurfer segmentation for the healthy control group and the COVID group. The COVID group shows increased χ in the brainstem, specifically in the Medulla and Pons (red arrows).

**Figure 2.**
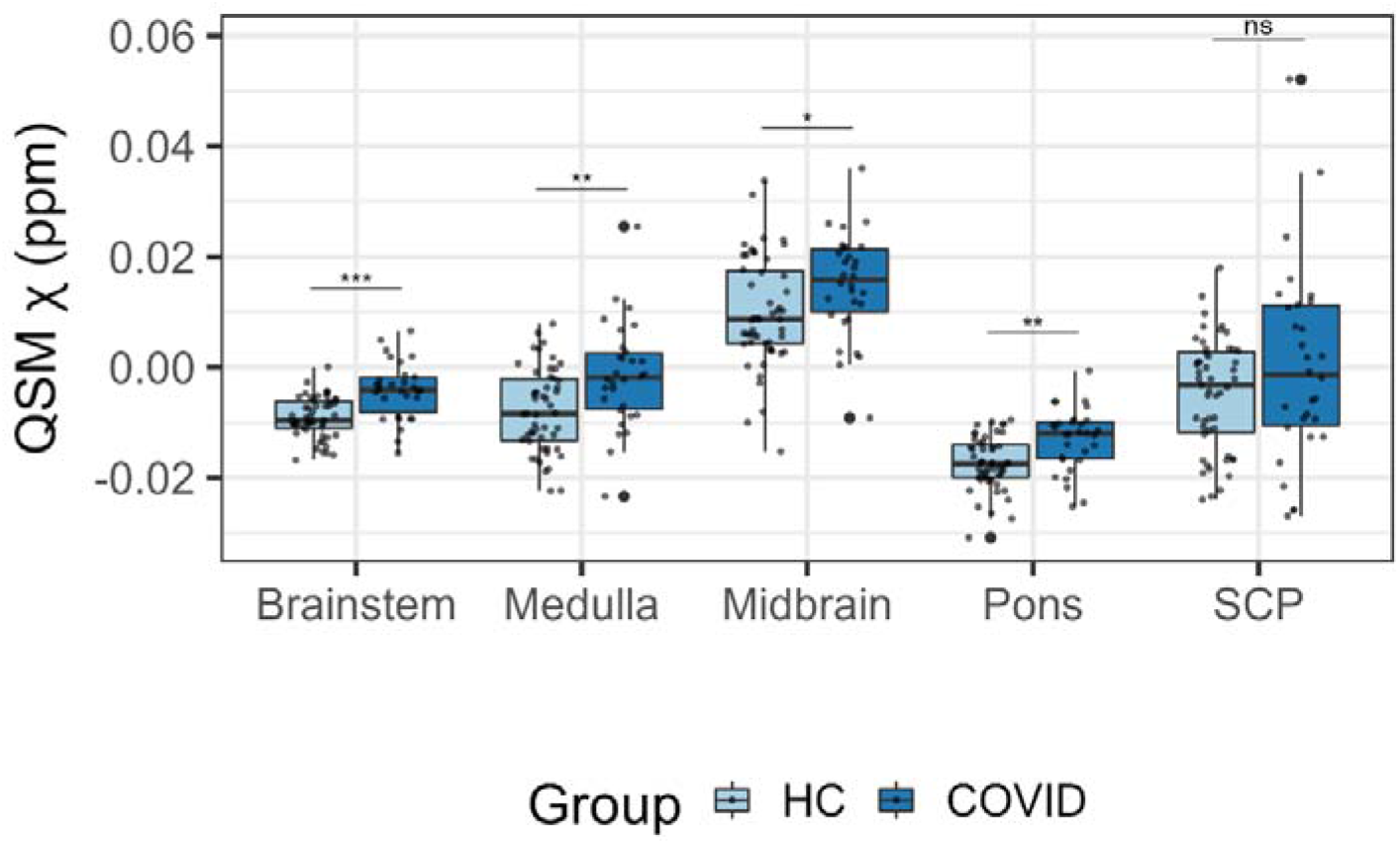
Boxplots of differences in the regional average χ between the COVID group and the HC group obtained from the Brainstem. Group differences assessed with a linear model with age, gender and age by gender interactions added as explanatory variables of no interest. FDR-corrected statistics represented on the boxplots. Legend: *** p<0.001, ** p<0.01, * p<0.05, ns=not significant, SCP=Superior Cerebellar Peduncle.

Voxel wise analyses on the brainstem identified two significant clusters located in the medulla region (Figure 3; Table 2) with significant increase in χ in the COVID group (Cluster 1: p(FDR) < 0.0001, d = 4.56, 95% CI [0.011, 0.028], Pr(post.) = 1.00, BF = 1073; Cluster 2: Cluster 1: p(FDR) < 0.0001, d = 4.86, 95% CI [0.0098, 0.023], Pr(post.) = 1.00, BF = 906). These clusters partially overlap brainstem regions known to be associated with respiratory function and body homeostasis, including the inferior medullary reticular formation nuclei, the raphe obscurus and pallidus. At the less stringent, but still significant, threshold corrected for multiple comparisons (FWE threshold p-value < 0.05) additional clusters were observed in the Pons and Midbrain sub-regions of the brainstem (Supplementary Information 7-9).

**Table 2.** Volume, maximum t-statistic, centre of gravity (COG), location in the brainstem, and overlapping Brainstem Navigator ROIs from the significant clusters determined with randomise function in FSL (TFCE corrected p<0.01, cluster inference t=2.5, cluster volume > 1 mm^3^). Clusters shown in Figure 3.

| Cluster # | Volume of cluster ($\text{mm}^3$) | Max t-statistic in Cluster | COG X | COG Y | COG Z | Location | Brainstem Navigator ROIs overlapping cluster | Brainstem Navigator ROI full description |
| --- | --- | --- | --- | --- | --- | --- | --- | --- |
| Cluster 1 | 96.75 | 5.83 | 179 | 167 | 28 | Medulla | iMRtl_r, iMRtm_l, iMRtm_r, iMRt_r, ROb, RPa | inferior medullary reticular formation (left and right), raphe obscurus, raphe pallidus. |
| Cluster 2 | 21.13 | 4.79 | 192 | 169 | 43.5 | Medulla | iMRt_l, iMRtl_l | inferior medullary reticular formation (left) |

**Figure 3.**
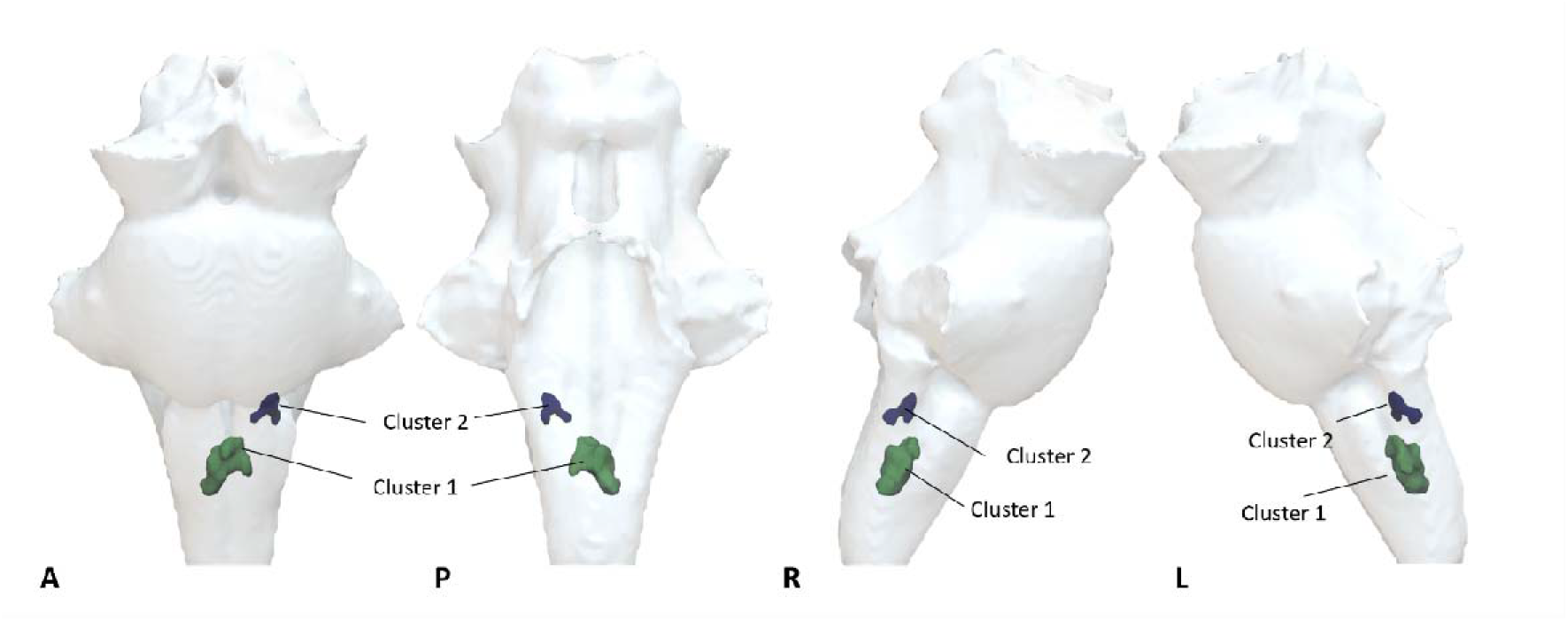
Voxelwise analysis showing increased QSM χ on the COVID group compared to HC. Significant clusters determined with randomise function in FSL (TFCE corrected p<0.01, cluster inference t=2.5, cluster volume > 1 mm^3^). (A) 3D projection on the brainstem ROI of the significant clusters.

### 3.2 Brainstem pathology and clinical assessments in patients

The mean χ extracted from the two clusters identified in the Medulla portion of the brainstem (FWE threshold p-value < 0.01) were positively associated with the highest CRP detected during admission (R = 0.36, p = 0.041, Pr(post.) = 0.84, BF = 5.1) and mRS (R = 0.60, p = 0.0046, Pr(post.) = 0.94, BF = 16.3). It was also weakly associated with the WHO severity index (R = 0.40, p = 0.046, Pr(post.) = 0.70, BF = 2.3) and period of hospital admission (R = 0.37, p = 0.054, Pr(post.) = 0.7, BF = 3.1) (Figure 4, Supplementary Information 10). There were no significant trends for other laboratory variables and clinical assessments (for highest D-Dimer during admission, GAD-7, PHQ-9 and lowest platelets during admission, R < 0.21, p > 0.15, Pr(post.) < 0.47 and BF < 0.88). No significant effect was found between the two clusters (for all variables, p > 0.60, Pr(post.) < 0.29, BF < 0.40) (Supplementary Information 10). Scatter plots (Figure 4), display the average χ extracted from the two clusters against the significant clinical outcome variables.

**Figure 4.**
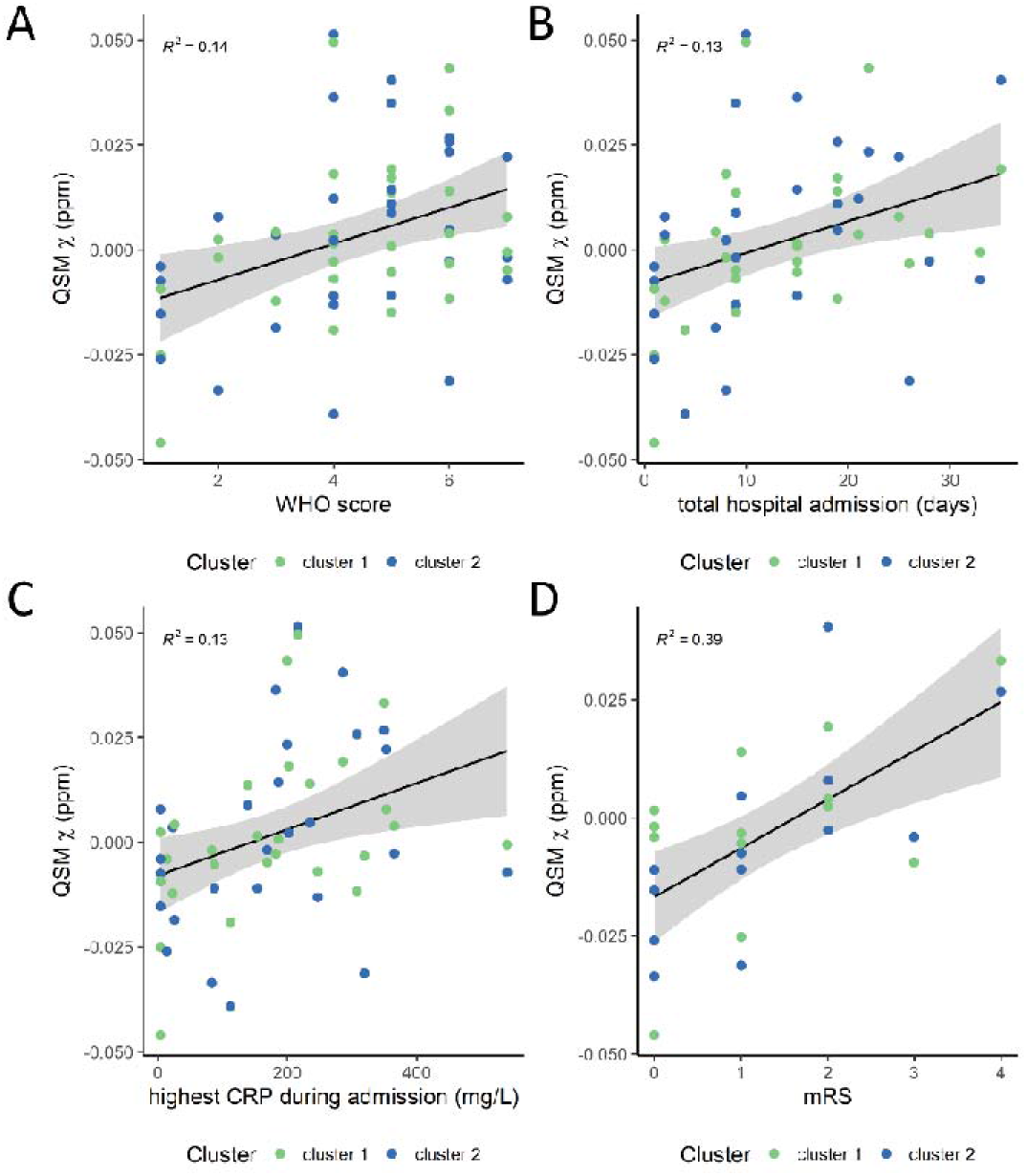
Scatter plots of the average QSM χ obtained on the clusters from the voxelwise group analysis with clinical and laboratory outcomes: (A) WHO score, (B) period of hospital admission, (C) highest CRP during admission, and (D) mRS. The R^2^ is also displayed in each plot. Plot (B) shows data without the outlier (Supplementary Information 5).

## 4. Discussion

The current study provides imaging evidence for mid-to-long term microstructural abnormalities in the brainstem following COVID-19 hospitalization. Our key findings are that in COVID-19 survivors, multiple regions of the medulla oblongata, pons and midbrain show magnetic resonance susceptibility abnormalities at a median time of 6.5 months from hospital admission. These differences are consistent with a neuroinflammatory response. These effects are independent of age and gender, and are more pronounced in those who had had more severe initial COVID-19 illness.

Symptoms of fatigue, dyspnea, breathlessness, cough and chest pain are common in the months after COVID-19 infection.^31–36^ Brainstem changes may predispose to, or exacerbate, such symptoms over and above peripheral organ damage. This role in the aetiology of long term symptoms may arise because the brainstem provides a nexus between sensory and motor inputs, and between the spinal cord and the brain, with nuclei that are responsible for controlling the sleep-wake cycle, respiratory drive, cardiac and vasomotor regulation. We hypothesise that a brainstem insult occurs in hospitalized COVID-19 patients, impairing autonomic functions that contribute to persisting clinical symptoms. In part, a similar pattern is observed following post severe traumatic brain injury, with patients reporting fatigue, dizziness, but also tachycardia, tachypnoea and hypertension^37,38^, linked to acute or chronic brainstem dysfunction.^39^

Neuropathological changes in the brainstem in patients with COVID-19 have been detected in *post mortem* cases.^11^ In most cases, there is no evidence of direct viral infection of the CNS, but rather a neuroinflammatory response to systemic infection. The process of increase in χ in patients recovering from COVID-19 infection is reminiscent of the observed inflammatory response in other neuroinflammatory disorders such as multiple-sclerosis.^40,41^ During acute inflammation, macrophage iron levels rise^42^, in concert with increased production of cytokines and reactive oxygen species.^43^ Indeed, increase of intracellular iron content can itself promote a proinflammatory state.^44^ These processes will tend to increase the paramagnetic properties of tissue, and, thereby increase tissue susceptibility as measured by QSM.

Our analysis was focused on the brainstem, exploring changes not only its sub-regions (midbrain, pons, medulla and SCP), but also on a voxel-by-voxel basis to allow increased anatomical resolution. The latter approach highlighted clusters in the inferior medullary reticular formation and in the raphe obscurus and pallidus, with increased tissue susceptibility in the COVID group compared to normal controls. The medullar reticular formation is responsible for the central control of the respiratory cycle. Nuclei included in the formation include the dorsal respiratory group (inspiratory centre) and the ventral respiratory group (with inhibitory and premotor expiration neurons).^45,46^ In addition, neurons in the raphe pallidus and obscurus have been found to be central chemoreceptors^46^ responsible for the full expression of ventilatory responses to hypercapnia.^47^ We propose that these changes provide evidence of a viral-induced proinflammatory state, which is responsible for impaired function in key brainstem circuits generating and controlling physiological allostasis.

CRP is a non-specific marker of inflammation or infection and has been found elevated in patients with COVID-19 and other acute respiratory syndromes such as the H1N1 influenza virus.^48^ Our results show that patients that had a greater peak inflammatory response during hospital admission (peak CRP), showed increased tissue susceptibility (likely associated with increased inflammation) in clusters within the medulla responsible for a regular autonomic respiratory function. In turn, patients with a more favourable functional outcome (modified Rankin Scale 0-2), with shorter hospital stays or lower COVID severity ratings showed decreased susceptibility in the medullary clusters. COVID-19 appears to drive a post-viral, long-lasting, hyperactivation of the immune system within the brainstem, impairing certain autonomic functions. In a similar manner, a portion of SARS and MERS survivors have shown similar long-lasting post-viral illnesses.^49–51^

In the brain, as first described by Raman *et al.*^52^ and later by Griffanti *et al.*^53^, susceptibility related changes in COVID-19 patients were found in the Thalamus in terms of T_2_* but not χ, which was attributed to differences in tissue compartmentalisation. In addition, Griffanti *et al.*^53^ found differences in χ in the right hippocampus. The authors argue that this could be related to higher iron accumulation related to virus infection, but could also be a partial volume issue of the MRI acquisition. An earlier analysis of a subset of our dataset^22^, did not show any QSM χ changes in these regions at 7T (which would be expected to enhance tissue susceptibility differences). Analysis of our full dataset consistently showed no group effects in the thalamus, hippocampus (Supplementary Information 11) or any other high-iron subcortical brain structures. However, our patients were scanned on average 219 days after hospital admission, which is over 3.5 times longer than timing of scans in these prior studies.^52,53^ Many brain changes normalize at 6-month follow-up imaging^54,55^ and these differences in scan timing could contribute to the difference in the results observed with our dataset.

Many studies have demonstrated that ultra-high field phase imaging improves contrast-to-noise ratio of cortical regions or iron-rich regions such as the globus pallidus or substantia nigra that have been used to assess changes in pathology.^21,56,57^ In this study we were able to highlight the importance of ultra-high field imaging to detect changes in the brainstem that were not previously reported. At 7T, QSM was able to detect negative and diffuse susceptibility values which were, on average across all healthy controls, -0.0091±0.0037 ppm. For COVID-19 patients there was nearly a 50% increase in the average χ to - 0.0042±0.0052ppm.

The study has several limitations. The sample size of patients is relatively small and heterogeneous. Recruitment of patients was challenging due to the contemporary safety concerns and lockdowns before widespread availability of vaccines. This study was a multi-centre effort. Our imaging results were indicative of negligible site effects for QSM providing increasing confidence on the applicability of T_2_* imaging for the CNS in multi-centre trials. We also acknowledge the gender imbalance of our normative dataset. The cohort was partly formed of data from a number of clinical studies acquired prior to the COVID-19 outbreak which contained a different gender balance. For this study we extracted a sample of healthy controls, selecting control cohorts where we were confident that the subjects had not experienced clinical or subclinical SARS-CoV-2 infection. In addition, all our group analyses were controlled for gender and age effects, and their interaction. For the voxelwise assessment, we used a conservative FWE threshold p-value of 0.01 for cluster inference and found two small clusters in the medulla region of the brainstem. This allowed us to isolate the most prominent peak locations that showed changes in our patient group compared to controls. At a lower threshold, other regions in the pons and midbrain showed increases in tissue susceptibility for the COVID patients, overlapping with the inferior olivary nucleus, the pontis oralis and caudalis, the ventral tegmental area, the periaqueductal gray and others (Supplementary Information 7). Future work utilizing brainstem MR susceptibility as a proxy of brain inflammation, together with further clinical indexes of sleep-wake cycle, cardiovascular and respiratory control metrics, might allow further understanding for which brainstem regions become impaired and to which extent. We also acknowledge that these scans were taken on a single timepoint after hospitalization (on average six months after hospitalization). Prospective follow-up studies would be helpful to understand the long-term sequelae of COVID-19 hospitalisation.

## 5. Conclusions

We show that the brainstem is a site of vulnerability to long term effects of COVID-19, with persistent changes evident in the months after hospitalization. These changes are hypothesised to be driven by neuroinflamatory responses, and more evident in patients with longer hospital stays, higher COVID severity, more prominent inflammatory responses, and worse functional outcomes. Ultra-high field 7T QSM was sensitive to these pathological changes in the brainstem, which could not be detected at standard clinical field strengths. This approach can provide a valuable tool to better probe the brain for the long-term effects of COVID-19 and other potential SARS-CoV diseases, in order to inform acute and long-term therapeutic strategies to aid recovery.

## 6. Data availability statement

We can provide average QSM χ extracted values from the brainstem and subregions upon reasonable request.

## 7. Acknowledgements

We thank our research participants for their invaluable dedication and inspiration. We thank the radiography, ethics and admin support teams at both sites for their involvement in the delivery of this project, especially developing new procedures for safe research during the COVID pandemic.

## 8. Funding

This work was supported by the NIHR Cambridge Biomedical Research Centre (BRC-1215-20014). The views expressed are those of the authors and not necessarily those of the NIHR or the Department of Health and Social Care. JBR was supported by the Wellcome Trust (103838; 220258) and Medical Research Council (M C_UU_00030/14). VFJN was supported by an Academy of Medical Sciences / The Health Foundation Clinician Scientist Fellowship. ETB was supported by an NIHR Senior Investigator award. BR, MC, SN are supported by NIHR Oxford BRC and BHF Oxford CRE. WTC was supported by funding from Wellcome Trust [225924/Z/22/Z]. CTR was supported by funding from Wellcome Trust [098436/Z/12/B]. For the purpose of open access, the authors have applied a CC-BY public copyright licence to any Author Accepted Manuscript version arising from this submission.

## 9. Competing interests

Dr. Pattinson is named as co-inventor on a provisional U.K. patent application titled “Use of cerebral nitric oxide donors in the assessment of the extent of brain dysfunction following injury. Dr Pattinson is named as co-inventors on a provisional U.K. patent titled “Discordant sensory stimulus in VR based exercise” UK Patent office application: 2204698.1 filing date 31/3/2022.

## Notes

### Author Declarations

The study was approved by the following ethics committees: Cambridgeshire Research Ethics Committee HBREC.2016.13.am3, East of England Research Ethics Committee 17/EE/0025 , Norfolk Research Ethics Committee EE/0395, and North West Preston Research Ethics Committee 20/NW/0235. All participants provided informed consent in accordance with the Declaration of Helsinki.

