## Supplementary File for "Quantitative susceptibility mapping at 7 Tesla in COVID-19: mechanistic and outcome associations"

### Supplementary Information


### C Rua et al.


*Supplementary Information 1 Demographics for the healthy controls (HC) and Covid+ subgroup data. Abbreviations: NC=data not collected, NA=not applicable, SpO2=blood saturation percentage, bpm=beat/breath per minute, GAD=Generalized Anxiety Disorder (Spitzer et al., 2006), PHQ=patient health questionnaire (Kroenke et al., 2001), mRS=modified Rankin Score* (Swieten et al., 1988).

| | HC | | COVID-19 | | |
|---|---|---|---|---|---|
| **Scanner Site** | **Site-1** | | **Site-1** | **Site-2** | **Statistical test (site-1 vs site-2)** |
| Subgroup | HC1 | HC2 | NA | | |
| N | 18 | 33 | 14 | 17 | NA |
| Age (y.o.) | 42±12 | 60±11 | 53±10 | 59±13 | p = 0.18 |
| Gender (male=M, female=F) | 18M,0F | 16M, 17F | 5M, 9F | 13M, 4F | ($\chi^2$ = 4.03, p = 0.13) |
| Period of hospital admission (days) | - | - | 21±34 | 13±9 | p = 0.36 |
| Time from admission to 7T MRI scan (days) | - | - | 195±33 | 214±107 | p = 0.20 |
| Highest CRP during admission (mg/L) | - | - | 157±144 | 207±132 | p = 0.34 |
| Highest D-Dimer during admission (ng/mL) | - | - | 1802±3047 | 8704±18970 | p = 0.19 |
| Lowest platelets during admission ($10^9$/L) | - | - | 223±49 | 179±43 | p = 0.018 |
| WHO severity scale (range: 0-10) | - | - | 3.7±2.2 | 4.9±1.3 | p = 0.081 |
| Breathlessness score (range: 0-10) | - | - | 2.8±2.4 | 1.1±1.8 | p = 0.11 |
| GAD-7 (range: 0-21) | - | - | 4.9±6.0 (range: 0-20) | 4.4±4.6 (range: 0-16) | p = 0.84 |
| PHQ-9 (range: 0-27) | - | - | 7.2±5.7 (range: 0-15) | 6.0±4.1 (range: 2-15) | p = 0.57 |
| mRS | - | - | 1.28±1.2 | NC | NA |

*Supplementary Information 2: Registration of the individual QSM maps to standard space*

In order to map each subject's χ maps to the 0.5mm isotropic ICBM 2009b standardized space for statistical analysis, the first echo of the $T_2^*$w imaging data was first registered to each subject's $T_1$w scan with ANTs. A one step registration was performed using a rigid registration at four convergence levels using cross-correlation for the similarity metric. Then, the $T_1$w images were mapped to standard space in three steps: first and second steps were performed each with four levels, with a mutual information metric, and with rigid and affine transformations, respectively; while the third step was performed with five convergence levels, a cross-correlation metric, and with the symmetric image normalization transformation method.

Because registration of the 3T $T_1$w data to standard space can be challenging for brainstem regions, an additional registration step was added to the three-step pipeline described above, using a target brainstem mask to limit the registration search area. For this registration, five convergence levels with a cross-correlation metric and the symmetric image normalization transformation method were used. The χ maps were then mapped to standard space using the registration estimates described above. χ maps were visually inspected in standard space for quality of registration both for the brain and brainstem regions.

*Supplementary Information 3: Differences across imaging site were tested on the COVID group. Boxplots split into site (site-1 (green) vs site-2 (orange)) showing differences in the regional average χ obtained from the Brainstem Atlas. FDR-corrected statistics represented on the boxplots. Legend: ns=not significant, SCP Superior Cerebellar Peduncle.*

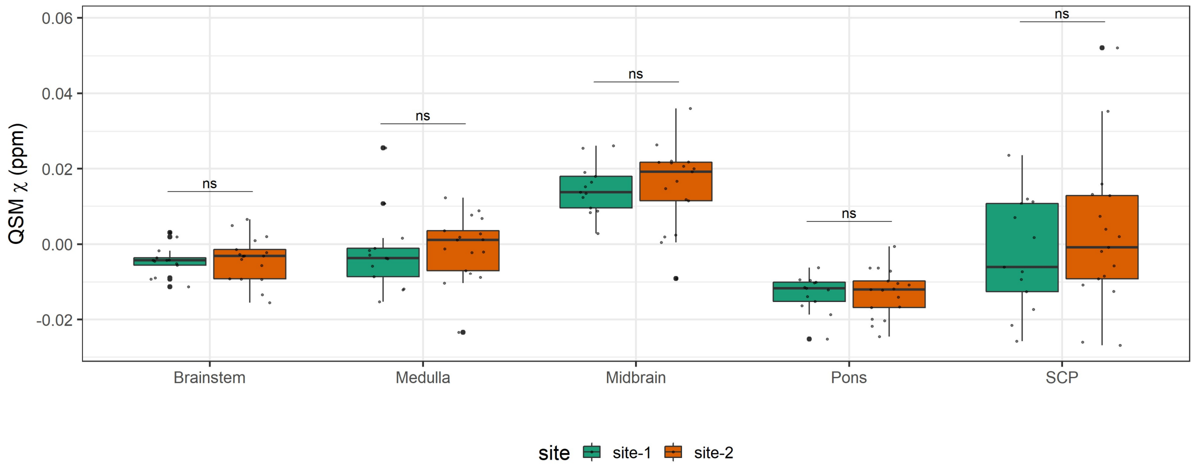


*Supplementary Information 4: Some clinical measurements collected from the COVID patients were not available. Therefore, linear mixed effects were performed by dropping patient data that did not include the clinical measurement of interest. Below is a table that describes the linear mixed effect model covariates and the number of subjects available for each test. Abbreviations: PerHospAdm= period of hospital admission, WHO=WHO severity scale, CRP= highest CRP during admission, DD=highest D-Dimer during admission, PLT=lowest platelets during admission, GAD-7= Generalised Anxiety Disorder-7 score, PHQ-9=Patient Health Questionnaire-9, mRS=modified Rankin Scale, TAdmtoScan= time from admission to 7T MRI scan, cluster_index=index of the cluster obtained from the randomise group analysis (subsection 2.4, main manuscript).*

| **Linear mixed effects model** | **Number of subjects(N)** |
|---|---|
| χ ~ PerHospAdm + age + gender + age*gender + TAdmtoScan + cluster_index | 31 |
| χ ~ WHO + age + gender + age*gender + TAdmtoScan + cluster_index | 31 |
| χ ~ CRP + age + gender + age*gender + TAdmtoScan + cluster_index | 28 |
| χ ~ DD + age + gender + age*gender + TAdmtoScan + cluster_index | 25 |
| χ ~ PLT + age + gender + age*gender + TAdmtoScan + cluster_index | 28 |
| χ ~ Breathlessness + age + gender + age*gender + TAdmtoScan + cluster_index | 21 |
| χ ~ GAD-7 + age + gender + age*gender + TAdmtoScan + cluster_index | 24 |
| χ ~ PHQ-9 + age + gender + age*gender + TAdmtoScan + cluster_index | 24 |
| χ ~ mRS + age + gender + age*gender + TAdmtoScan + cluster_index | 14 |

*Supplementary Information 5: Scatter plots of the average QSM χ obtained on the clusters from the voxelwise group analysis with the period of hospital admission, (A) including all subjects, (B) excluding outlier – subject with period hospital admission = 134 days. The $R^2$ is also displayed in each plot.*

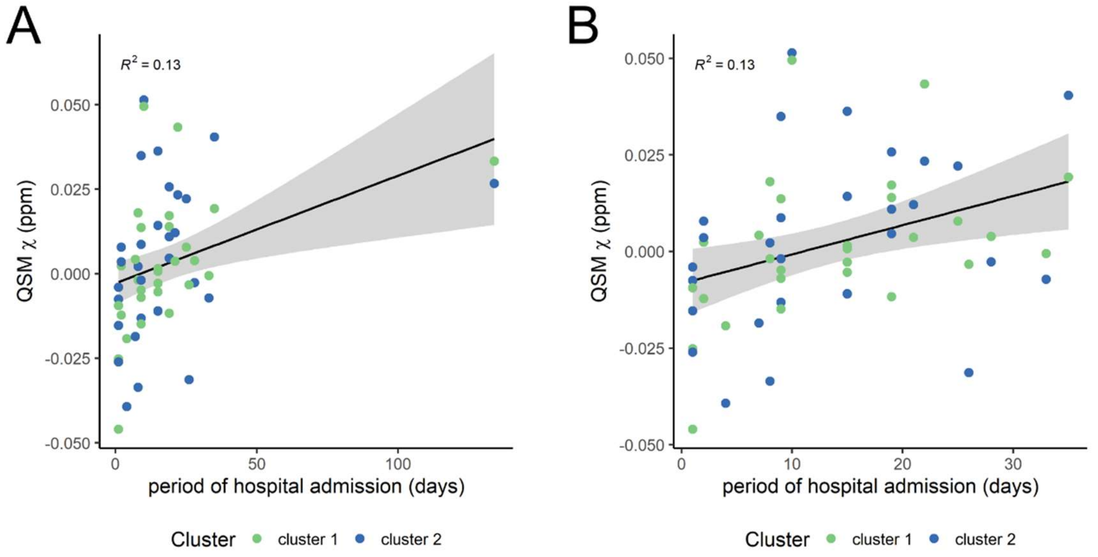


*Supplementary Information 6: (A) QSM (B) phase and (C) magnitude data from COVID patient showing lack of viable MR signal in the brainstem. Range of QSM map -0.1 to 0.1 ppm. Range of phase map -3.14 to 3.14 radians, range of magnitude map 0 to 702.*

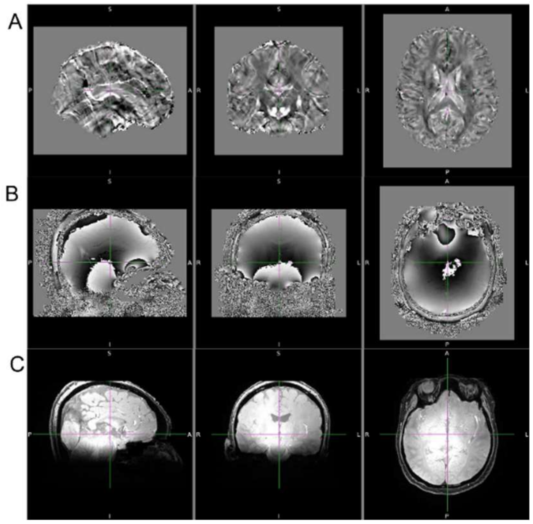

*Supplementary Information 7: Cluster regions showing significant increase in QSM $\chi$ in the COVID group compared to the healthy control group (TFCE corrected $p<0.05$, cluster threshold $t=2.5$). Legend: COG=center of gravity. Brainstem Navigator ROIs that overlapped the significant clusters described in the last two columns.*

| Cluster # | Volume of cluster [$mm^3$] | Max t-statistic in Cluster | COG X | COG Y | COG Z | p (FDR) | Cohen's d | 95% CI [ppm] | Pr(posterior) | BF | Location | Brainstem Navigator overlapping ROIs | Brainstem Navigator ROI full description |
|---|---|---|---|---|---|---|---|---|---|---|---|---|---|
| C1 | 390.25 | 5.83 | 182 | 168 | 34.5 | <0.0001 | 5.0 | [0.0097 0.023] | 1.00 | 2556 | Medulla | iMRt_l, iMRt_r, iMRtl_l, iMRtl_r, iMRtm_l, iMRtm_r, ION_r, ROb, RPa, sMRt_l, sMRtl_l, Ve_l, VSM_l, VSM_r | inferior medullary reticular formation (left and right), inferior olivary nucleus (right), raphe obscurus, raphe pallidus, superior medullary reticular formation (left), vestibular nuclei complex (left), viscero-sensory-motor nuclei complex (left and right) |
| C2 | 297 | 4.68 | 180 | 180 | 58.4 | <0.0001 | 5.3 | [0.010 0.023] | 1.00 | 22863 | Pons, Medulla | ION_r, LDTg_CGPn_r, PCRtA_l, PnO_PnC_l, RPa, Ve_r | inferior olivary nucleus (right), laterodorsal tegmental nucleus – central gray of the rhomboencephalon, paravicellular reticular nucleus - alpha part (left), pontis oralis and caudalis (left), raphe pallidus, vestibular nuclei complex (right) |
| C3 | 105.5 | 4.58 | 191 | 219 | 75.7 | <0.0001 | 5.6 | [0.022 0.047] | 1.00 | 7108 | Pons | - | |
| C4 | 102.5 | 4.9 | 203 | 206 | 94 | <0.0001 | 4.9 | [0.012 0.029] | 1.00 | 3632 | Pons, Midbrain | - | |
| C5 | 48.875 | 4.42 | 189 | 193 | 80.3 | <0.0001 | 4.4 | [0.0083 0.022] | 1.00 | 773 | Pons | - | |
| C6 | 20.875 | 5.21 | 188 | 188 | 39.3 | <0.0001 | 4.7 | [0.013 0.033] | 1.00 | 2069 | Medulla | - | |
| C7 | 16.5 | 3.85 | 183 | 211 | 59.9 | <0.0001 | 3.7 | [0.014 0.047] | 0.98 | 61 | Pons | - | |
| C8 | 14.75 | 4.38 | 162 | 191 | 92.2 | <0.0001 | 4.2 | [0.0095 0.027] | 1.00 | 349 | Pons | - | |
| C9 | 10.625 | 4.29 | 215 | 199 | 79.6 | 0.0017 | 3.2 | [0.0083 0.036] | 0.97 | 34 | Pons | - | |
| C10 | 7.75 | 4.08 | 157 | 222 | 103 | 0.00027 | 3.9 | [0.018 0.056] | 0.99 | 148 | Midbrain | - | |
| C11 | 4 | 3.84 | 170 | 222 | 79.9 | 0.0022 | 3.3 | [0.014 0.058] | 0.95 | 20 | Pons | - | |
| C12 | 3.625 | 4.84 | 174 | 222 | 123 | <0.0001 | 5.0 | [0.022 0.051] | 1.00 | 7271 | Midbrain | VTA_PBP_r | ventral tegmental area-parabrachial pigmented nucleus complex (right) |
| C13 | 2.5 | 4.79 | 173 | 190 | 136 | <0.0001 | 4.3 | [0.017 0.046] | 1.00 | 997 | Midbrain | SC_r | superior colliculus (right) |
| C14 | 2 | 3.99 | 167 | 159 | 17.3 | 0.00020 | 4.1 | [0.010 0.029] | 1.00 | 209 | Medulla | - | |
| C15 | 1.625 | 3.38 | 173 | 216 | 72.2 | 0.0023 | 3.1 | [0.0089 0.041] | 0.96 | 23 | Pons | - | |
| C16 | 1.625 | 3.23 | 201 | 220 | 91.4 | 0.0045 | 3.1 | [0.0095 0.044] | 0.91 | 10 | Pons | - | |
| C17 | 1.5 | 3.92 | 162 | 227 | 107 | 0.000039 | 4.6 | [0.020 0.050] | 1.00 | 1753 | Midbrain | PAG | periaqueductal gray |
| C18 | 1.5 | 4.16 | 178 | 193 | 134 | 0.00043 | 3.6 | [0.017 0.058] | 0.99 | 106 | Midbrain | - | |
| C19 | 1 | 3.96 | 170 | 209 | 52.6 | 0.00043 | 3.9 | [0.014 0.042] | 0.99 | 90 | Pons | - | |

*Supplementary Information 8: 3D projection on the brainstem ROI of the voxelwise analysis showing increased QSM χ on the COVID group compared to healthy controls. Significant clusters determined with randomise function in FSL (TFCE corrected p < 0.05, cluster inference t = 2.5, cluster volume > 1 mm³).*

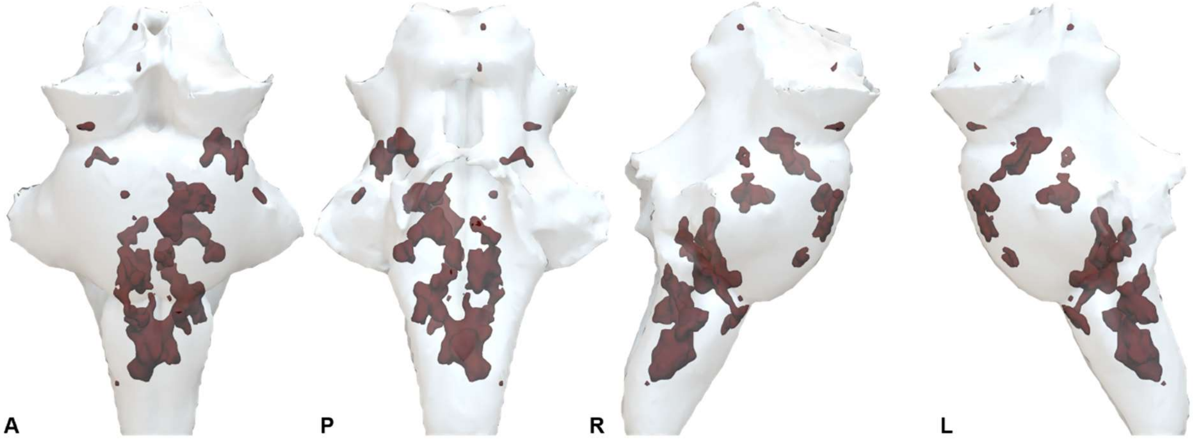


*Supplementary Information 9: Overlay of t-statistic scores of the brainstem COVID-vs-HC group voxelwise analysis on the MNI 2009b brain. The results for the FWE corrected p-value < 0.01 (top row) and p-value < 0.05 (bottom row) are displayed for reference.*

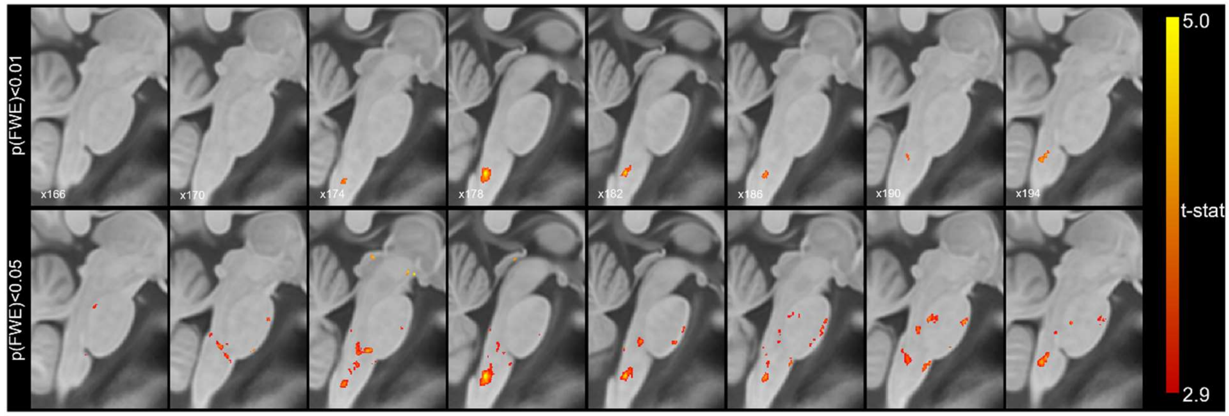

*Supplementary Information 10: Table displaying the correlation coefficient (R), p-value, p-posterior and Bayes factor (BF) for the correlation statistics of the QSM χ averaged in the brainstem clusters and the clinical and laboratory outcomes. Abbreviations of fixed effect variables: PerHospAdm= period of hospital admission, WHO=WHO severity scale, CRP= highest CRP during admission, DD=highest D-Dimer during admission, PLT=lowest platelets during admission, GAD-7= Generalised Anxiety Disorder-7 score, PHQ-9=Patient Health Questionnaire-9, mRS=modified Rankin Scale.*

| | | **WHO** | **PerHospAdm** | **PerHospAdm without outlier** | **CRP** | **DD** | **GAD-7** | **PHQ-9** | **PLT** | **mRS** |
|---|---|---|---|---|---|---|---|---|---|---|
| **Correlation coefficient, R** | main fixed effect | 0.40 | 0.35 | 0.37 | 0.36 | 0.21 | 0.15 | -0.071 | 0.11 | 0.60 |
| **lmer, p-value** | main fixed effect | 0.046 | 0.025 | 0.054 | 0.041 | 0.15 | 0.65 | 0.94 | 0.51 | 0.0046 |
| | ROI effect | 0.90 | 0.90 | 0.86 | 0.77 | 0.84 | 0.66 | 0.66 | 0.77 | 0.60 |
| **p-posterior** | main fixed effect | 0.70 | 0.86 | 0.76 | 0.84 | 0.47 | 0.28 | 0.25 | 0.34 | 0.94 |
| | ROI effect | 0.21 | 0.21 | 0.22 | 0.22 | 0.23 | 0.23 | 0.23 | 0.22 | 0.29 |
| **BF** | main fixed effect | 2.3 | 6.1 | 3.1 | 5.13 | 0.88 | 0.39 | 0.34 | 0.52 | 16.3 |
| | ROI effect | 0.27 | 0.27 | 0.28 | 0.28 | 0.29 | 0.30 | 0.30 | 0.28 | 0.40 |

*Supplementary Information 11: Boxplots of differences in the regional average χ between the COVID group and the healthy control (HC) group obtained from the Harvard-Oxford Atlas. Group differences assessed with a linear model with age and gender, and age by gender interaction added as explanatory variables of no interest. FDR-corrected statistics represented on the boxplots. Legend: *** p<0.001, ** p<0.01, * p<0.05, ns not significant, CSF=cerebral spinal fluid, GM=grey matter, WM=white matter.*

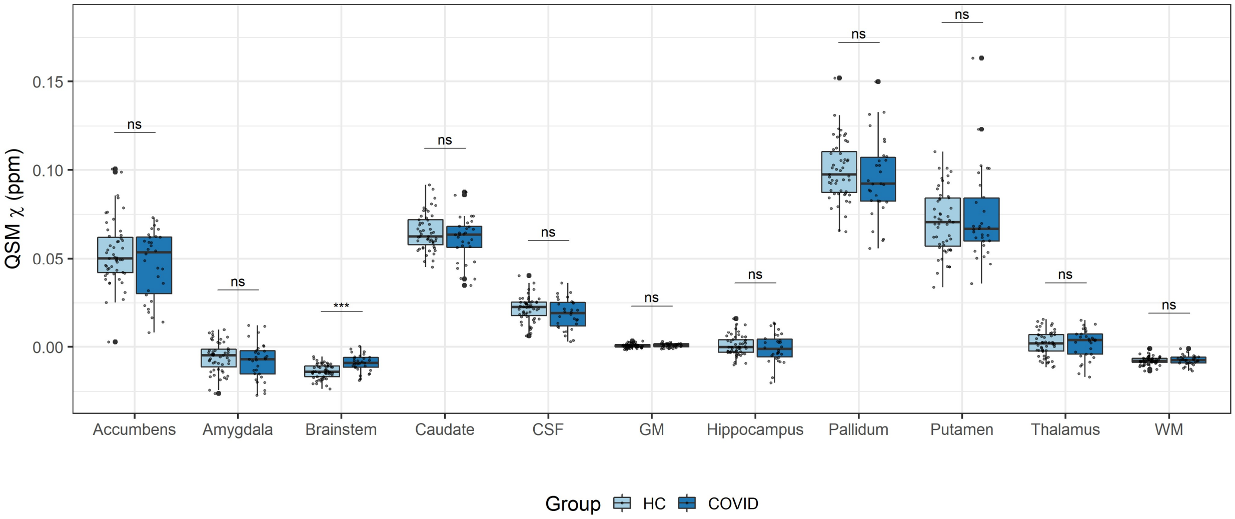
